# Inequalities in COVID-19 inequalities research: who had the capacity to respond?

**DOI:** 10.1101/2021.09.27.21264156

**Authors:** Joan Benach, Alvaro Padilla, Lucinda Cash-Gibson, Diego F. Rojas-Gualdrón, Juan Fernández-Gracia, Víctor M. Eguíluz, the COVID-SHINE group

## Abstract

The COVID-19 pandemic has been testing countries’ capacities and scientific preparedness to actively respond, and collaborate on a common cause. It has also heightened awareness of the urgent need to empirically describe and analyse health inequalities, to be able to act effectively. What is known about the rapidly emerging COVID-19 inequalities research field? We analysed the volume of COVID-19 inequalities scientific production (2020-2021), its distribution by country income groups and world regions, and inter-country collaborations, to provide a first snapshot. COVID-19 inequalities research has been highly collaborative, however inequalities exist within this field, and new dynamics have emerged in comparison to the global health inequalities research field. To ensure preparedness for future crises, investment in health inequalities research capacities must be a priority for all.

## Introduction

Since the start of the COVID-19 pandemic social inequalities have been exacerbated, there has been a heightened awareness of the need to empirically describe and analyse health inequalities (1, 2), and report on context-specific actions to effectively address them (2, 3). In many ways, the COVID-19 pandemic can be interpreted as a sort of natural experiment that has been testing countries’ capacities and scientific preparedness to actively and effectively respond to such crises, to gather and comprehensively analyse health and socio-demographic data, and to develop comprehensive equity-oriented COVID-19 analyses (3), as well as collaborate, at local, national and global levels for a common cause.

Prior to the pandemic, evidence demonstrated that inequalities existed in this type of research capacity globally (4), which has likely hindered many countries’ research capacity and preparedness to respond to the COVID-19 pandemic (3). At present, many research questions appear: What do we know about the emerging COVID-19 inequalities research field? What research patterns and trends have emerged so far? What is being produced, where, and indirectly where are the capacity and scientific knowledge gaps? Which countries are collaborating together on this important topic?

By analysing the volume of global scientific production on COVID-19 inequalities over the past year (2020-2021), the distribution of this scientific production by country income groups and world regions, and the inter-country collaborations within this scientific production, this study aims to provide a first snapshot of this rapidly emerging research field.

## Methods

### Design

We conducted a bibliometric analysis and network analysis of original scientific articles on COVID-19 inequalities, published between January 2020 and April 23^rd^ 2021.

### Search strategy

Scientific articles were retrieved from Scopus database, as this allows for bibliometric analysis and provides a larger access to articles than other databases such as Web of Science (5). Our search strategy combined terms to identify scientific articles on both COVID-19 and health inequalities, building on a previous bibliometric analysis on health inequalities research (4). We exported the metadata, including the citation and bibliographical information of the documents, their abstracts, keywords, and references. The following search formula was used: [Title-abstract-keyword]: ((covid* PRE/3 *19) OR (*19 PRE/3 covid*) OR (coronavirus PRE/3 *19) OR (*19 PRE/3 coronavirus) OR (“novel coronavirus”) OR (“new coronavirus”) OR (“SARS-CoV-2”) OR (“2019-nCoV”)) AND ((inequ*) OR (dispari*) OR (equit*) OR (equal*) OR (povert*) OR (*igual*) OR (“pobreza”) OR (equid*) OR (*soci* PRE/2 gradient) OR (“gender”) OR (“género”) OR (ethni*) OR (“race”) OR (“racism”) OR (social PRE/2 class) OR (prejud*) OR (marginal*) OR (refugee*) OR (migrant*) OR (“homeless”) OR (“housing”) OR (“precarious”) OR (unemploy*))

### Variables

We calculated bibliometric variables by country, according to authors’ country of affiliation: the number of co-authored articles, the number of correspondence author articles, and the number of scientific collaborations for each possible pair of countries. To standardize research output by country size, we calculated rates per country population: the Country Research Production (CRP) rate is defined as the number of co-authored articles per million habitants. The country correspondence author rate is defined as the number of correspondence authors per million habitants. We obtained the most recent estimation of the population from the World Bank Database (6). From this source, we additionally extracted classifications for income groups and world regions.

### Data analysis

Bibliometric analyses were performed with the Bibliometrix 3.1.3 (7) package for R 4.1.0. Statistical analyses were performed in Stata 17.0.

### Bibliometric and network analysis

We describe the research output, the average citations per year per article, the average number of authors per article, and the collaboration index: the average number of authors per article considering only multi-authored articles. The landscape of international collaborations in COVID-19 inequalities research was studied by a joint analysis of the research output and collaborations based on Louvain’s clustering algorithm only considering countries with a minimum of 10 collaborations with other countries.

### Statistical analysis

The differences in COVID-19 inequalities research between countries, according to income group and world region, were quantified with productivity rate ratios (RR). The generalized linear model with Poisson distribution and logarithmic link function was used to estimate RR and 95% confidence intervals (95% CI).

### Network analysis and inequalities

For each nation we computed the Gini coefficient for the distribution of articles in collaboration with other countries. The analysis was performed using Python. The value of the Gini coefficient is shown as a function of the number of collaborators in Fig. 2.

## Results

We found 9,355 documents in 2,588 sources. 140 countries contributed at least one article. A total of 43,713 authors contributed to global scientific production on COVID-19 inequalities. The average citation per document was 8.7. The three highest country producers were: the United States (US) (32% of the total scientific production), the United Kingdom (UK) (10%), and China (6%).

The contribution to this research field was highly unequal between countries of different income groups and world regions (Figure 1). High-income countries showed a CRP rate 33 times higher and a correspondence author rate 54 times higher than low income countries. These differences were smaller, though still substantial, for upper middle income and lower middle income countries, whose CRP rates were six and three times higher, and correspondence author rates were nine and two times higher, respectively, than that of low income countries. As for the inequalities between world regions, in comparison to Sub-Saharan Africa, North America’s CRP rate and correspondence author rate were 39 and 59 times higher, Europe’s were 12 and 23 times higher, the Middle East & North Africa’s were seven and 12 times higher, East Asia & the Pacific’s were seven and ten times higher, South Asia’s were two and three times higher, and Latin America & Caribbean’s were two times higher.

**Fig. 1.**
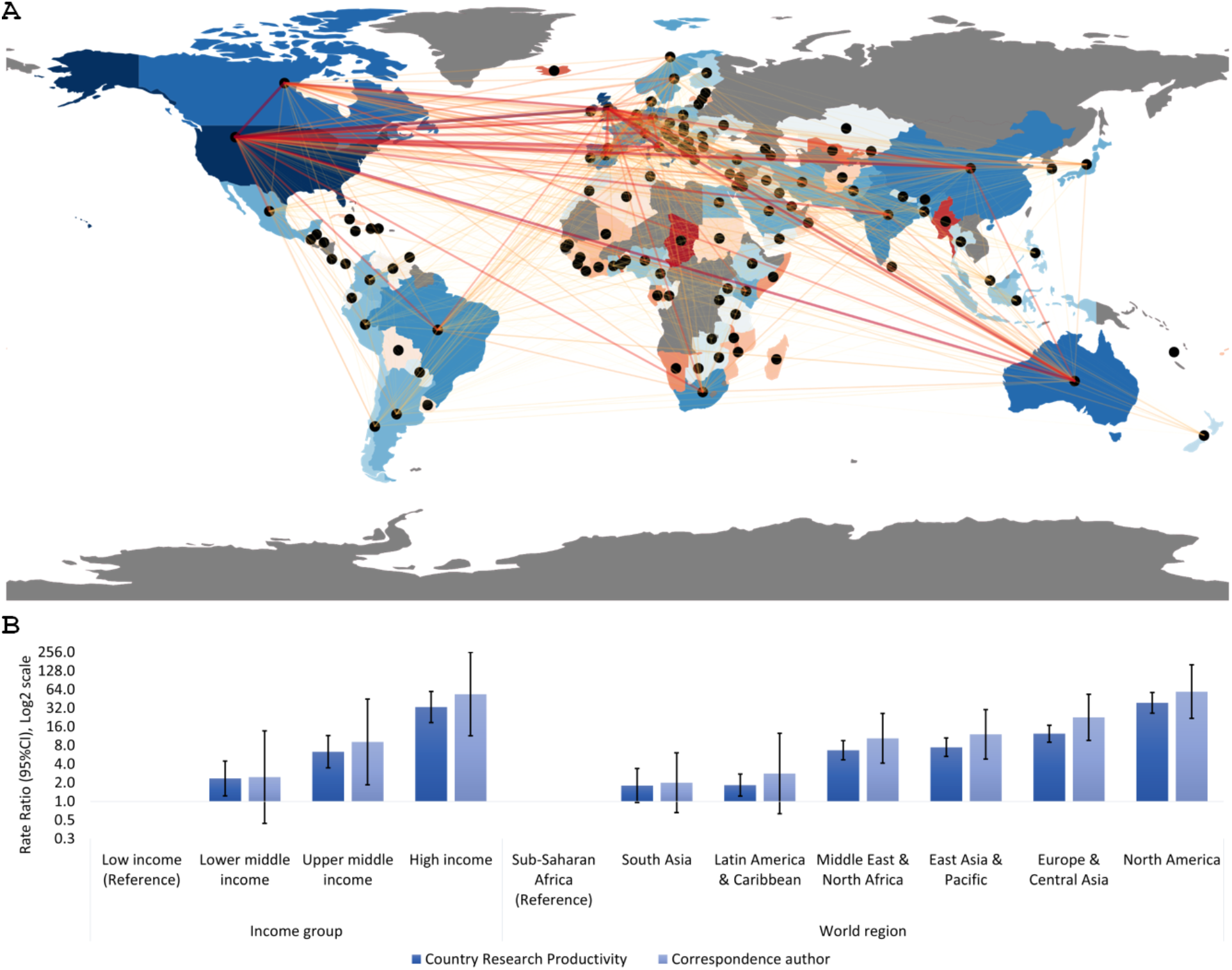
Inequalities in COVID-19 inequalities research production. **(A)** Map of the world showing the different collaborations between countries. The color of the country reflects the number of articles in collaboration with other countries (increasing from red to blue). The color of the connections encodes the number of articles in which the two linked countries appear. **(B)** Country Research Productivity and Correspondence author rates per million people by country income group and world region.

COVID-19 inequalities scientific production has been highly collaborative, 84.5% of the articles (n=7902) had multiple co-authors, and had a collaboration index of 5.5. Nevertheless, inequalities were also present in those collaborations (Figure 2). High income countries had the largest number of international collaborators. In addition, as the total number of collaborators increased, so did the Gini coefficient between collaborators. The presence of some lower middle income country outliers such as Kenya is explained by their participation in a large amount of research, which was led by authors affiliated to institutions of higher income groups.

**Fig. 2.**
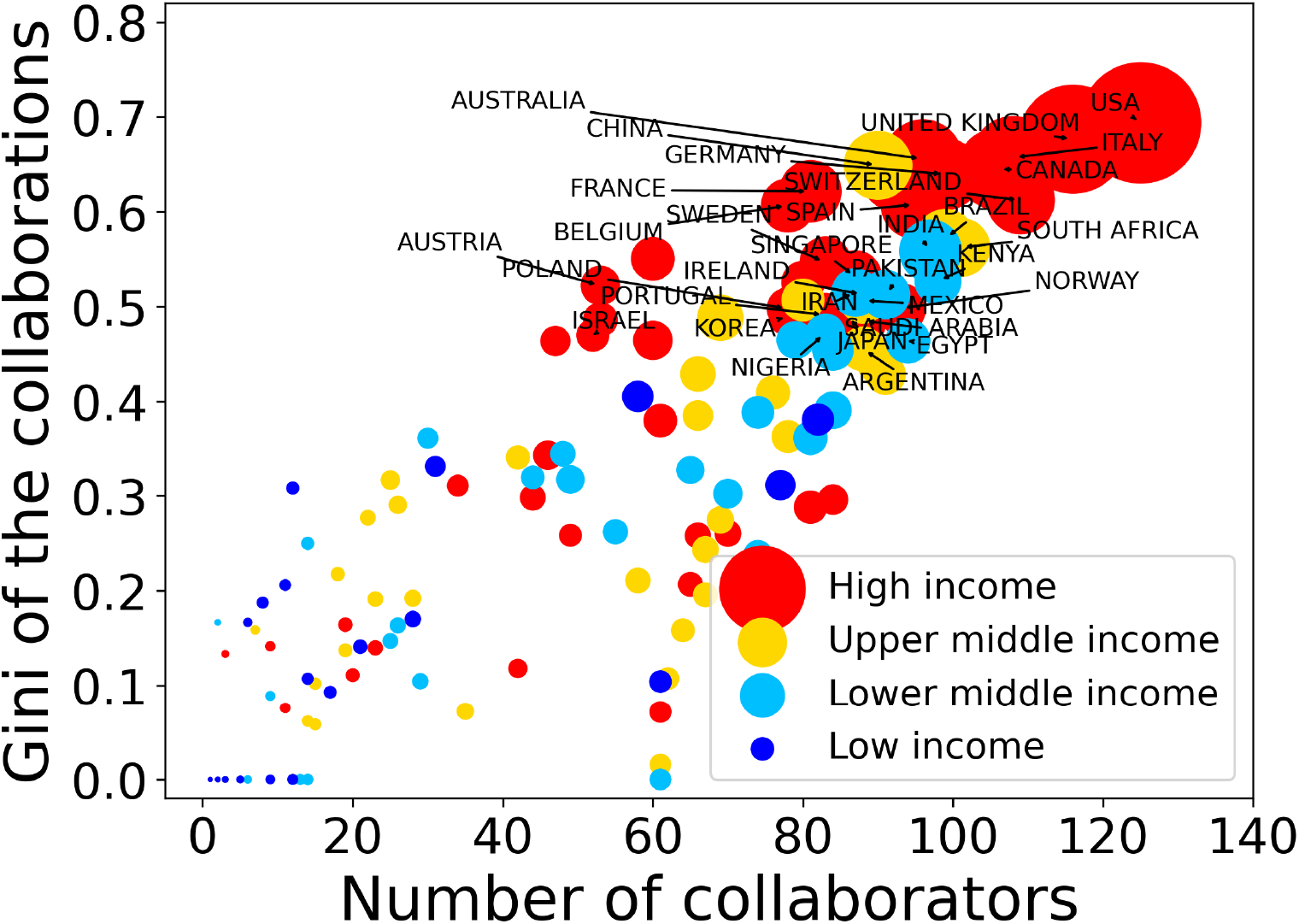
Gini coefficient between international collaborators, as a function of the number of collaborators per nation. Size of the symbols reflects the number of articles for each nation in collaboration with other nations. Color indicates the income group of the nation. Only the names of the 30 nations with higher Gini and number of collaborating nations are shown.

Three main collaboration network clusters were identified (Figure 3). One (green) cluster was formed mainly by the Anglosaxon, Asian and African countries. A second (red) cluster was formed mainly by European countries, and Chile. A third (blue) cluster was formed by Colombia, Brazil, Mexico and Argentina. The US and the UK had the most predominant presence in the global collaborations map, and were also the two countries with the highest scientific production.

**Fig. 3.**
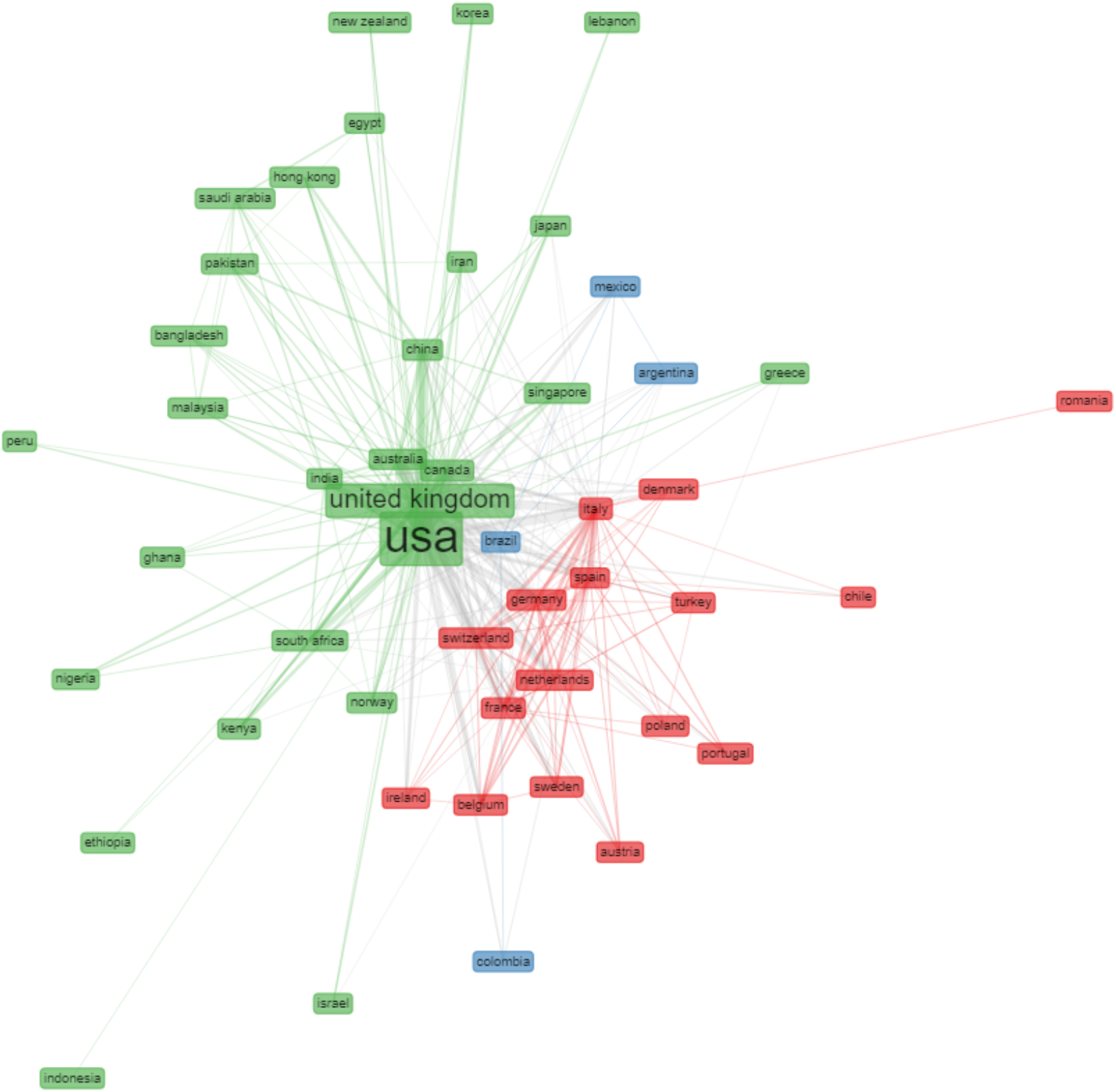
Collaboration network cluster for countries with a minimum of 10 collaborations. Each color represents a cluster, and the size of each rectangle represents countries’ total production of COVID-19 inequalities research.

## Discussion

The COVID-19 inequalities research field has rapidly emerged and expanded over the past year, yet we have identified notable inequalities in research productivity rates between countries. We identified the US, UK and China as the three highest country producers of COVID-19 inequalities research, which is in line with the results of the recent bibliometric analysis of global scientific research on COVID-19 (8). Many of our results also concur with the findings from a previous historical and global bibliometric analysis of the health inequalities research field (1966-2015) (4), which identified inequalities within the health inequalities research field and collaborative practices. For example, the US and UK are the most predominant in these research collaborations, and are the two countries which have produced and co-produced the most scientific research on this topic; also there is still a notable European research collaboration cluster.

Interestingly though, in comparison with this previous bibliometric analysis (4), a number of differences have emerged in the COVID-19 inequalities research patterns and trends. Firstly, the US is at the center of all the collaborative networks in this research field, followed by the UK, as opposed to the UK being at the centre (4). Secondly, a new (blue) cluster has emerged consisting of Colombia, Brazil, Mexico and Argentina. These four countries were previously identified as the four highest producers of health inequalities research in the Latin American and Caribbean region (1966-2015) (4), and are known to have had a tradition of collective health and social medicine research, indicating strong health inequalities research capacity (9). Curiously, Chile does not form part of this cluster, although it seems to have retained strong collaborations with Brazil, over the past year. Rather, Chile forms part of the predominantly “European” (red) cluster, this may partly be due to the fact that there were pre-existing collaborations between research groups within Spain and Chile, for example, around so-called “…s*ocio-critical accounts of work-related and migrant health” (p*.*5)* (10), which may likely have been leveraged during the pandemic.

Lastly, the Middle Eastern and North African region has a high productivity rate and correspondence author rate, and is almost on par with the East Asian and the Pacific region, as the third largest regional producer of COVID-19 inequalities research. In comparison to the previous analysis of the health inequalities research field (4), the Middle Eastern and North African region’s scientific production has increased rapidly over the past few years, surpassing that of Latin America and Caribbean. In addition, previously the region had its own distinct health inequalities research collaboration cluster (4); however, for COVID-19 inequalities research, several countries from the region form part of the predominantly “Anglosaxon” (green) cluster. This finding may be connected to the fact that in 2019, a new Commission on Social Determinants of Health was established in the WHO Eastern Mediterranean region, led by Sir Michael Marmot and his team at the Institute of Health Equity, University College London, UK, and in collaboration with the World Health Organization’s Alliance for Health Policy and Systems Research. The Commission objectives have been to analyse health inequalities across the region, “…*build knowledge and evidence for action”*, (p.5)(11), and develop context-specific recommendations to reduce health inequalities across the region. As such, subsequent research collaborations on COVID-19 and inequalities may have been established, building on these newly formed collaborations.

The COVID-19 pandemic has been testing countries’, communities and individuals in many ways, particularly in terms of their capacities and preparedness to actively and effectively respond to such a global crisis, and to collaborate to manage it. This capacity, and the type of policy response taken are, however, conditioned by a number of pre-existing contextual factors, such as the level of pre-existing health inequalities, politics, ideology and value perspectives on health inequalities, and what each society considers socio-politically desirable (e.g. collectivism versus individualism) (3, 12, 13). In addition to a country’s economic resilience, public trust in government, and general trust in science (3, 14, 15, 16). Furthermore, while research collaborations can help to mobilise and share resources, which can help to strengthen research capacities and bring mutual benefits (12), they also bring new conditions, interests and power relations to the research production process (12).

While there has been a bibliometric analysis on the global COVID-19 research field more broadly (8), to the best of our knowledge this is the first bibliometric and network analysis of the COVID-19 inequalities research field more specifically. In terms of study limitations, while our results are based only on empirical articles published in international academic journals, they provide a useful overview of the global dynamic and patterns within this newly emerging scientific field, and pose many interesting research questions that require further exploration. Further research is needed to expand on these findings, to establish more in depth understanding of why and how these trends and outcomes might have occurred, as well as the type of COVID-19 inequalities research that has been produced in different countries (3), as this may indicate priority areas for action. In addition, it would be important to assess the relationship between productivity research rates by country and other variables of interest, such as the share of the GDP invested in research and development, to further explore the determinants of these research capacities.

Collectively, what is clear is the need for every country to develop stronger capacity to collect timely, reliable, disaggregated by different social groups (e.g. social class, age, gender, ethnicity/race, geography), to establish comprehensive COVID-19 data collection systems to be able to report and monitor on health inequalities (3, 11, 12, 17, 18, 19, 20). In addition, ethical principles must be instilled in future research collaborations to ensure more equitable partnerships (12, 21, 22). Conducting rapid, and comprehensive COVID-19 inequalities analyses can assist to produce vital knowledge about COVID-19 related incidences, eaths, and social inequalities in health (3, 12, 23, 24). This knowledge is needed to help to guide the development of more evidence-informed policy and action, and raise general awareness at the local, national, and global level. In order to support this, and ensure better preparedness for future crises, investment into public health and health inequalities research capacities should be a priority for all.

## Data Availability

All the data used for this paper is available upon request.

